# Demographic and Comorbidity Characteristics of U.S. Veterans in a Retrospective Descriptive Matched Cohort Study Addressing Problematic Gambling

**DOI:** 10.64898/2026.10.30.26366936

**Authors:** T. Elizabeth Workman, Joel Kupersmith, Christopher Spevak, Friedhelm Sandbrink, Qing Treitler-Zeng

## Abstract

**Introduction:** Problematic gambling is an increasing public health concern that also affects U.S. Veterans. Problematic gambling in Veterans correlates with several other factors, including socio-economic issues like homelessness and legal problems, and mental health issues like post-traumatic stress disorder, and suicide attempt. Our objective was to analyze differences between U.S. Veterans experiencing problematic gambling compared to similar U.S. Veterans who did not, according to diagnostic codes.

**Materials and Methods:** Electronic health records from the U.S. Department of Veterans Affairs (VA) for health care encounters between 2016-01-01 and 2024-12-31 served as data for this study. We performed a one-to-one match on age, race, ethnicity, gender, and VA station for 775 Veterans diagnosed with problematic gambling by International Classification of Diseases 10^th^ edition codes, to Veterans not receiving one of the diagnostic codes relevant to gambling. We then compared comorbidity diagnoses, marital status, and VA care encounters, applying chi-squared and t-test analyses.

**Results:** There were many statistically significant differences in care use, marital status, and comorbidities. Veterans with problematic gambling were more likely to experience other substance abuse, including opioid use disorder (22.5% vs 8.4%) and alcohol use disorder (56.3% vs 25.9%). Veterans with problematic gambling were also more likely to experience mental illness, including major depressive disorder (64.9% vs 46.7%), post-traumatic stress disorder (50.8% vs 43.1%), and suicide attempt (5.3% vs 3.0%), and socio-economic issues, including economic hardship (28.7% vs 16.3%), homelessness (33.9% vs 14.8%), legal problems (13.4% vs 6.8%) and social/family problems (53.6% vs 46.5%). Veterans experiencing problematic gambling also were less likely to be married and received more VA outpatient care.

**Conclusions:** The findings suggest that problematic gambling in Veterans correlates with worse outcomes when comparing similar Veterans receiving care through the same facilities. It is feasible to screen for problematic gambling in primary care encounters, however the extent of screening in VA care is unknown. Screening for problematic gambling in VA primary care may enable providers to assist patients experiencing problematic gambling to find directed care.

## Introduction

Problematic gambling is an increasing, significant public health concern [1]. According to the International classification of diseases 10^th^ edition (ICD 10), it is defined as “frequent, repeated episodes that dominate the patient’s life to the detriment of social, occupational, material, and family values and commitments” [2]. L. T. Tran, et al. found that worldwide, 46.2% of adults and 17.9% of adolescents had gambled in the last year, with approximately 11.9% of men and 5.5% of women having experienced some type of risk gambling, equating to approximately 439.6 million adults engaged in any risk gambling globally [1]. This has been fueled by online gambling [3] including the increasing popularity of gambling apps [3]. Risk factors for problematic gambling include male gender[4-7], young age [6-8], living alone [4], being single [4, 6] or married for less than five years [4], impulsivity [9], and high stress levels [10]. Problematic gambling is associated with several comorbidities, including schizophrenia [11], depression [11, 12] bipolar disorder [11], anxiety [13], group B personality disorder [11], other substance use disorders [13, 14], and relationship and financial problems. [14] Suicide is a major concern in gambling, with one in five individuals with problematic gambling attempting suicide in their lifetime [15-18].

Current and past U.S. military personnel are also susceptible to problematic gambling. Active-duty service members are at particular risk due to factors such as age, military culture, and accessibility [19]. The National Council on Problem Gambling (NCPG) reports that military personnel are 2-3 times more likely to experience problematic gambling, with as many as 56,000 active duty service members meeting criteria for gambling disorder [20]. Statistics are incomplete due to the hesitancy to self-report as well as challenges with systematic screens [21]. As with other substance use disorders, problematic gambling may result in readiness issues for the fighting force. Current efforts are underway in the U.S. Department of Defense to promote education, diagnosis and treatment [22]. Studies estimate that nearly 9% of Veterans experience clinical lifetime problematic gambling [18, 23]. Problematic gambling is also associated with many comorbidities in Veterans, including mental health issues such as other substance abuse [24], mood disorders [25], including major depression [26], obsessive-compulsive disorders [27], antisocial behavior disorder [28], post-traumatic stress disorder [29, 30], and anxiety [31]. Veterans experiencing problematic gambling may also have suicide ideation [32]. Kausch found that 39.5% of 114 Veteran compulsive gamblers in a gambling treatment program had attempted suicide at some point in life [33]. Problematic gambling issues for Veterans also correlate with pain [32] and prior trauma (emotional, physical, or sexual abuse) [34]. Socio-economic issues also correlate with problematic gambling in Veterans, including homelessness [35], less social support [30], and legal issues [35]. Information is limited, lacking comparisons of Veterans with problematic gambling to matched counterparts.

Screening for gambling in primary care and similar environments is feasible [36]. The extent of screening for problematic gambling in Veterans Affairs (VA) care is unknown; Etuk et al. noted a lack of standardized screening across U.S. Federal agencies serving military and post-military individuals [18].

Problematic gambling can be recorded using ICD diagnostic codes. How do Veterans who are diagnosed with problematic gambling compare to similar Veterans who are not recorded as receiving the diagnosis? We compared comorbidities, marital status, and care use among Veterans diagnosed with problematic gambling matched to other Veterans on age, race, ethnicity, gender, and VA station. A station is a service region within the VA that typically includes one or more hospitals, and other care facilities such as outpatient clinics. Our objective was to analyze differences between U.S. Veterans experiencing problematic gambling compared to similar U.S. Veterans who did not, according to ICD 10 diagnostic codes. All research was performed in the VA Informatics and Computing (VINCI) secure platform [20]. To our knowledge, this is the first nation-wide study of its kind to compare U.S. Veterans diagnosed with problematic gambling by ICD code to matched similar U.S. Veterans.

## Materials and Methods

This was a retrospective descriptive matched cohort study. We used a national database of electronic health records (EHRs) of Veterans from previously reported research [37] as a data source. Our study cohort consisted of 366,089 distinct patients who received outpatient care between 2016-01-01 and 2024-12-31. We identified patients with at least one relevant ICD 10 code (F63.0- or Z72.6-) during a VA outpatient encounter between 2016-01-01 and 2024-12-31. For the patients identified as having a relevant ICD 10 code we performed a one-to-one ratio match for age, race, ethnicity, VA station, and gender, on patients not receiving one of the relevant ICD 10 codes within the same time period. In this study, the patients having received one of the relevant ICD 10 codes is referred to as the problematic gambling group (PGG) and the matched patients as “the matched patients”. By matching on age, race, ethnicity, VA station, and gender, we maximized the amount of unique one-to-one matches of patients with access to the same general care, i.e. receiving care from the same VA station.

We then determined which patients had received one or more of several subsequent comorbidity diagnoses, using the first gambling diagnosis date as an index date for each matched pair. To determine comorbidity diagnosis, we used ICD 10 code sets established by several VA research groups and care facilities as described in prior research [37]. For “other victimization” we did not include Y07 codes that indicate crime perpetration. Marital status was also retrieved for all patients. Outpatient encounter counts on or after the index date were also recorded, with a maximum of one encounter per day counted. We then ran chi-squared tests, using SAS, version 9.4, and the t-test on outpatient encounters using the scipy.stats python library.

## Results

Of 366,089 total patients having an outpatient VA visit between 2016-01-01 and 2024-12-31, 839 patients received at least one relevant ICD 10 code (Figure 1). We were able to perform a unique one-to-one match on age, race, ethnicity, VA station, and gender, on patients not receiving a relevant ICD 10 code within the same time period (i.e., matched patients) for 775 of the patients receiving a relevant ICD 10 code (i.e., PGG patients). Of the 775 matched patient pairs, there were 101 stations represented.

**Figure 1.**
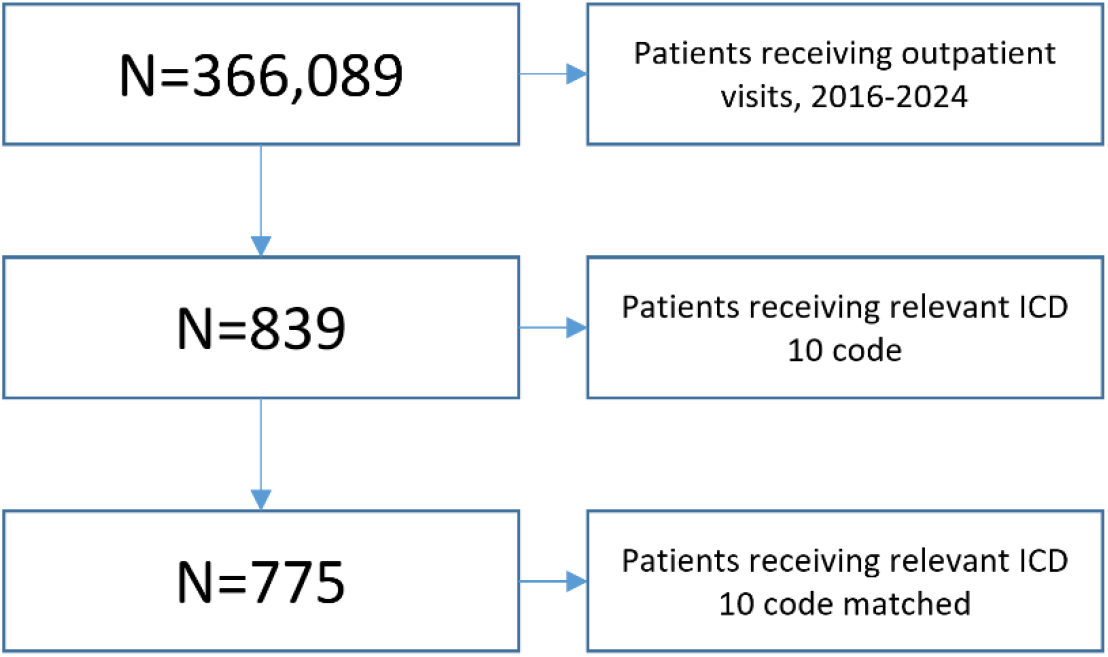
Cohort creation.

Demographic data on matched patient pairs are in Table 1. The average age at index date was 56.4 years of age, with ages ranging from 20.0 years of age to 92.8 years of age. The majority of patients were White or Black/African American.

**Table 1.**
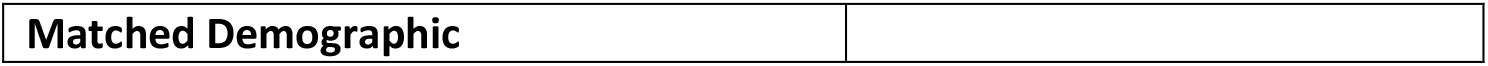
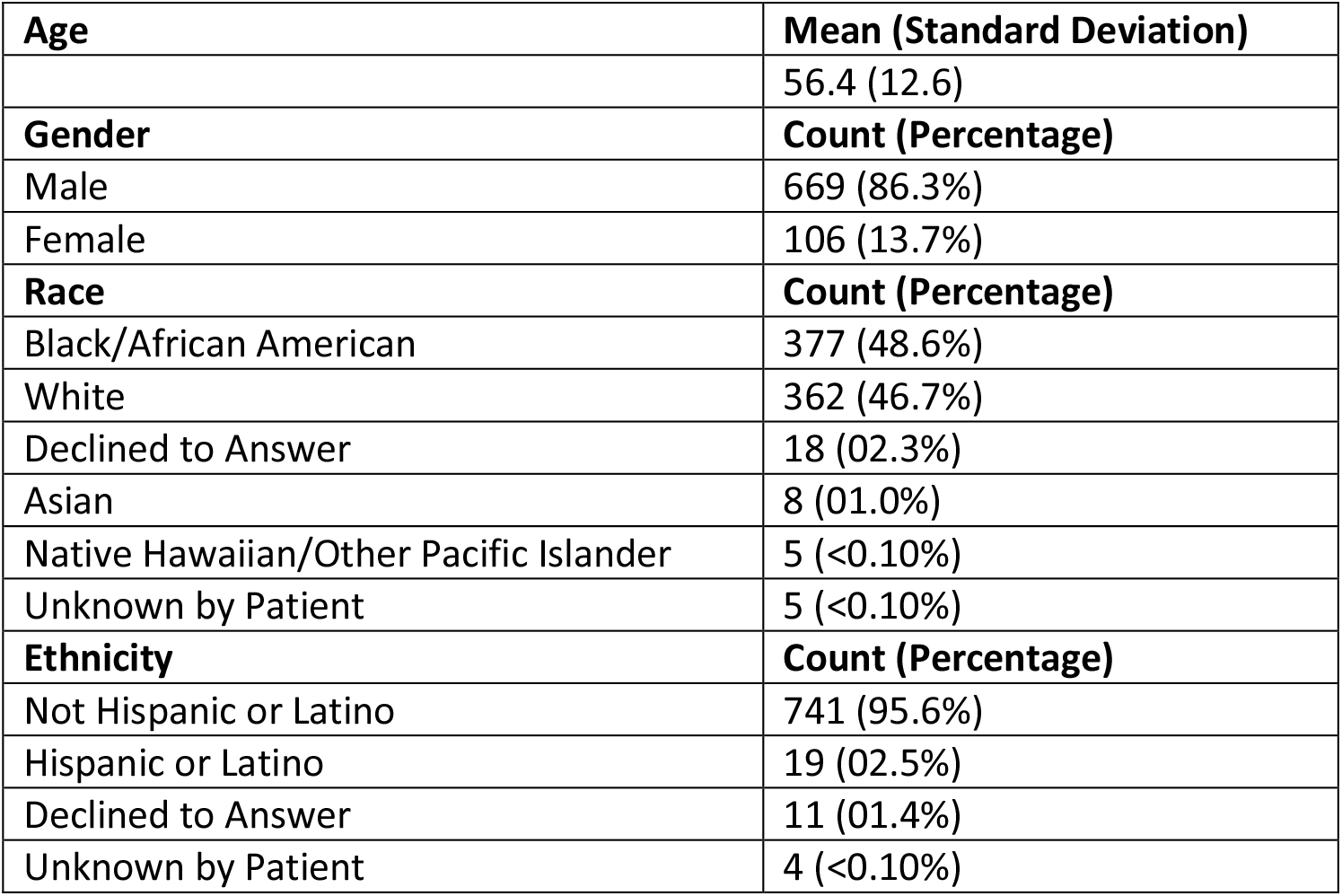
Demographic data on matched pairs.

Comorbidity, marital status, and care encounter results are in Table 2. Significant results are in bold text at a 0.05 alpha threshold (red: PGG; blue: matched patients; green: marital status of all patients).

**Table 2.** Comorbidities, Marital Status, and outpatient encounters. Items and values significant for PGG patients in bold red; items and values significant for matched patients in bold blue; Marital Status in bold green.

| **N** | **775** | **775** |  |
|---|---|---|---|
| **Comorbidity** | **PGG** | **Matched Patients** | **X<sup>2</sup> p-value** |
| Acute Myocardial Infarction | 35 (4.5%) | 29 (3.7%) | 0.4437 |
| Abdominal & Bowel Pain | 302 (39%) | 295 (38.1%) | 0.7148 |
| Anorexia | 1 (0.13%) | 1 (0.13%) | 1.000 |
| Anxiety | 122 (15.7%) | 106 (13.7%) | 0.2512 |
| **Any Malignancy** | **100 (12.9%)** | **128 (16.5%)** | **0.0447** |
| **Alcohol Use Disorder** | **436 (56.3%)** | **201 (25.9%)** | **<.0001** |
| Back Pain | 411 (53%) | 401 (51.7%) | 0.611 |
| Bulimia | 0 (0%) | 1 (0.13%) | 0.3173 |
| Cerebrovascular Disease | 123 (15.9%) | 138 (17.8%) | 0.3086 |
| **Congestive Heart Failure** | **129 (16.7%)** | **167 (21.6%)** | **0.0141** |
| Chronic Hepatitis B | 10 (1.3%) | 9 (1.2%) | 0.8174 |
| Chronic Pulmonary Disease | 264 (34.1%) | 242 (31.2%) | 0.2334 |
| Chronic Kidney Disease | 86 (11.1%) | 110 (14.2%) | 0.0666 |
| **Chronic Obstructive Pulmonary Disease** | **208 (26.8%)** | **168 (21.7%)** | **0.0178** |
| Dementia | 59 (7.6%) | 48 (6.2%) | 0.2704 |
| Diabetes | 325 (41.94%) | 290 (37.4%) | 0.0692 |
| **Drug Abuse Dependence Psychosis** | **380 (49%)** | **191 (24.7%)** | **<.0001** |
| **Economic Hardship** | **222 (28.7%)** | **126 (16.3%)** | **<.0001** |
| **Hepatitis C** | **88 (11.4%)** | **54 (7%)** | **0.0028** |
| **Heart Failure** | **114 (14.7%)** | **151 (19.5%)** | **0.0126** |
| **Homelessness** | **263 (33.9%)** | **115 (14.8%)** | **<.0001** |
| Hypertension | 540 (70%) | 536 (69.2%) | 0.8255 |
| **Insomnia** | **246 (31.7%)** | **203 (26.2%)** | **0.016** |
| Ischemic Stroke | 55 (7.1%) | 60 (7.7%) | 0.628 |
| **Legal Problems** | **104 (13.4%)** | **53 (6.8%)** | **<.0001** |
| **Major Depressive Disorder** | **503 (64.9%)** | **362 (46.7%)** | **<.0001** |
| Neck Pain | 246 (31.7%) | 250 (32.3%) | 0.8276 |
| Nightmares | 20 (2.6%) | 18 (2.3%) | 0.7426 |
| Obesity | 304 (39.2%) | 330 (42.6%) | 0.1792 |
| **Opioid Overdose** | **9 (1.2%)** | **0 (0%)** | **0.0026** |
| **Other Victimization** | **118 (15.2%)** | **87 (11.2%)** | **0.0201** |
| **Opioid Use Disorder** | **174 (22.5%)** | **65 (8.4%)** | **<.0001** |
| **Post-traumatic Stress Disorder** | **394 (50.8%)** | **334 (43.1%)** | **0.0023** |
| **Schizophrenia & Schizoaffective Disorder** | **104 (13.4%)** | **61 (7.9%)** | **0.0004** |
| Sexually Transmitted Infection | 49 (6.3%) | 45 (5.8%) | 0.6703 |
| **Social/Family Problems** | **415 (53.6%)** | **360 (46.5%)** | **0.0052** |
| **Suicide Attempts** | **41 (5.3%)** | **23 (3.0%)** | **0.0216** |
| TBI | 27 (3.5%) | 15 (1.9%) | 0.0605 |
| **Marital Status** | **PGG** | **Matched Patients** | **X<sup>2</sup> p-value:<br>0.0002** |
| Married | 227 | 305 |  |
| Divorced | 251 | 219 |  |
| Never Married | 203 | 174 |  |
| Separated | 64 | 43 |  |
| Widowed | 29 | 27 |  |
| Unknown/Missing | 1 | 7 |  |
| **Average Outpatient Care Encounters (standard deviation)** | **341 (297.56)** | **286 (272.39)** | **t-test p-value:<br>0.0001** |

The PGG patients had significantly higher rates of other substance use-oriented conditions (opioid use disorder, opioid overdose, alcohol use disorder, drug abuse dependence psychosis), mental health issues (major depressive disorder, post-traumatic stress disorder, schizophrenia and schizoaffective disorder, suicide attempts), socio-economic issues (economic hardship, homelessness, legal problems, other victimization, social/family problems) as well as chronic obstructive pulmonary disease, hepatitis C, and insomnia. The matched patients experienced higher rates of any malignancy, congestive heart failure, and heart failure. PGG patients had significantly more care visits in the VA. PGG patients were also more likely to not be married.

## Discussion

There were multiple differences between the PGG patients and the matched patients, and many of the findings from this study correlate with previous research. PGG patients were more likely to experience other substance use issues, mental health issues, and socio-economic issues including homelessness. The PGG group was also less likely to be married, which may correlate with less social support. [30] There was a significant increase in suicide attempts (Table 2), with 5.3% of the PGG patients having received a reported suicide attempt (recorded by ICD 10 code) versus 3.0% in the matched group. This compares with a reported lifetime suicide attempt of one out of 5 individuals with problematic gambling [15-18], and 39.5% of 114 Veteran compulsive gamblers attempting suicide at some point in life [33]. This study captured suicide attempt in a comparably smaller timeframe. Also, the patients in the cited studies [15-18, 33] were in treatment for, or otherwise proactively seeking help for problematic gambling, therefore it may be possible they were more willing to report suicide issues than the patients in this study. It is unknown if the patients in this study were receiving treatment for problematic gambling. Also, in the patient groups in this study, it is possible that suicide attempt is underreported, because recorded suicide attempts were perhaps known only because the patient was treated for the suicide attempt at a VA facility.

It is unknown how many VA patients experience problematic gambling, because the extent of screening for it is unknown. As before noted, earlier research has documented a lack of screening [18] and problematic gambling may be underreported by patients. The number of patients identified in this study by ICD diagnostic codes may represent a small subset of VA patients experiencing problematic gambling. Many of the PGG patients in the study also experienced other addictive behaviors (Table 2). It is possible that problematic gambling may have come up during conversations regarding other addictions with VA healthcare providers for these patients. The higher rates of COPD in the PGG patients may be associated with smoking, and the higher rates of hepatitis C in the PGG patients may be associated with drug misuse. More research is needed to explore these potential links.

Overall, these findings suggest a correlation between Veterans experiencing problematic gambling and worse outcomes when compared to other similar Veterans receiving care through the same facilities. Screening for problematic gambling in VA primary care may help identify Veterans with problematic gambling, so that they might receive directed care for this issue. These results further strengthen the need to examine this topic in both policy and research areas within the Veterans Health System. Because problematic gambling has been merged with substance use disorders in the Diagnostic and Statistical Manual of Mental Disorders V [38], particular efforts to screen substance use disorder patients for problematic gambling is warranted. If a Veteran is experiencing problematic gambling, a provider could seek if financial assistance is needed for treatment to potentially avoid added financial burden to the Veteran.

### Limitations

Because this study was dependent on ICD 10 diagnostic codes to identify problematic gambling, and implementation of screening for it in the VA is unknown, our research may provide information on only a portion of Veterans experiencing problematic gambling. Also, because of a potential lack of screening for all patients, it is possible that some patients in the matched group also experience problematic gambling. The VA serves a population largely consisting of older males; thus these findings may have limited generalizability to the world or U.S. population as a whole. However, the population the VA serves is evolving to include more females and younger patients. This is a descriptive study, and findings should not be interpreted as causal.

### Future Work

This work is part of a larger study addressing substance abuse and care in Veterans [37]. To understand problematic gambling in Veterans, it is paramount to understand screening practices and VA clinicians’ viewpoints regarding it. Future work will address this need. Future work may also include a natural language processing analysis of VA EHR clinical notes for problematic gambling documentation.

## Conclusion

Problematic gambling is a growing public health issue. It affects many people worldwide, including Veterans of the U.S. military. This study compares Veterans diagnosed with problematic gambling by ICD 10 code to similar Veterans not diagnosed with problematic gambling. The findings indicate that Veterans diagnosed with problematic gambling are significantly more likely to experience other substance abuse issues, socio-economic issues, and mental health issues, including suicide attempts. Screening practices for problematic gambling in the VA are uncertain; screening for problematic gambling in VA primary care may identify patients at risk and enable relevant care.

## Acknowledgments

We thank the United States Department of Veterans Affairs for funding this work. The views expressed are ours, the authors, and do not necessarily reflect those of the Department of Veterans Affairs, the United States Government, or the affiliated academic institutions.

## Data Availability Statement

The data used in the analysis include protected health information (PHI). Therefore, they are not publicly available.

## Institutional Review Board

This research was approved by the VA IRBNet protocol #1607134, date of approval: 26 March 2021.

## Ethics of Investigation

Studies involving humans were approved by The Washington DC VA Medical Center (Washington, DC). All research was conducted in accordance with local legislation and institutional requirements. Written informed consent was not required. No patients were contacted, only data from the EHR database were used.

## References

[1] L. T. Tran et al., “The prevalence of gambling and problematic gambling: a systematic review and meta-analysis,” Lancet Public Health, vol. 9, no. 8, pp. e594–e613, Aug 2024, doi: 10.1016/S2468-2667(24)00126-9.

[2] W. H. Organization, “International Classification of Diseases 10th edition, Version:2019.” [Online]. Available: https://icd.who.int/browse10/2019/en (accessed 30 Oct 2026).

[3] E. G. Deans, S. L. Thomas, M. Daube, and J. Derevensky, ““I can sit on the beach and punt through my mobile phone”: The influence of physical and online environments on the gambling risk behaviours of young men,” Soc Sci Med, vol. 166, pp. 110–119, Oct 2016, doi: 10.1016/j.socscimed.2016.08.017.

[4] M. Cakici, E. Cakici, A. Babayigit, and M. Karaaziz, “Gambling Behaviour: Prevalence, Risk Factors and Relation with Acculturation in 2007-2018 North Cyprus Adult Household Surveys,” J Gambl Stud, vol. 37, no. 4, pp. 1099–1111, Dec 2021, doi: 10.1007/s10899-021-10008-3.

[5] C. Cavalera et al., “Italian Adult Gambling Behavior: At Risk and Problem Gambler Profiles,” J Gambl Stud, vol. 34, no. 3, pp. 647–657, Sep 2018, doi: 10.1007/s10899-017-9729-8.

[6] N. Hing, A. Russell, B. Tolchard, and L. Nower, “Risk Factors for Gambling Problems: An Analysis by Gender,” J Gambl Stud, vol. 32, no. 2, pp. 511–34, Jun 2016, doi: 10.1007/s10899-015-9548-8.

[7] N. Hing, A. M. Russell, and M. Browne, “Risk Factors for Gambling Problems on Online Electronic Gaming Machines, Race Betting and Sports Betting,” Front Psychol, vol. 8, p. 779, 2017, doi: 10.3389/fpsyg.2017.00779.

[8] S. Buth, F. M. Wurst, N. Thon, H. Lahusen, and J. Kalke, “Comparative Analysis of Potential Risk Factors for at-Risk Gambling, Problem Gambling and Gambling Disorder among Current Gamblers-Results of the Austrian Representative Survey 2015,” Front Psychol, vol. 8, p. 2188, 2017, doi: 10.3389/fpsyg.2017.02188.

[9] G. Florez, P. A. Saiz, E. M. Santamaria, S. Alvarez, L. Nogueiras, and M. Arrojo, “Impulsivity, implicit attitudes and explicit cognitions, and alcohol dependence as predictors of pathological gambling,” Psychiatry Res, vol. 245, pp. 392–397, Nov 30 2016, doi: 10.1016/j.psychres.2016.08.039.

[10] D. F. K. Wong, X. Y. Zhuang, A. Jackson, N. Dowling, and H. H. M. Lo, “Negative Mood States or Dysfunctional Cognitions: Their Independent and Interactional Effects in Influencing Severity of Gambling Among Chinese Problem Gamblers in Hong Kong,” J Gambl Stud, vol. 34, no. 3, pp. 631–645, Sep 2018, doi: 10.1007/s10899-017-9714-2.

[11] A. Bergamini et al., “At-risk gambling in patients with severe mental illness: Prevalence and associated features,” J Behav Addict, vol. 7, no. 2, pp. 348–354, Jun 1 2018, doi: 10.1556/2006.7.2018.47.

[12] R. A. Volberg, L. M. McNamara, and K. L. Carris, “Risk Factors for Problem Gambling in California: Demographics, Comorbidities and Gambling Participation,” J Gambl Stud, vol. 34, no. 2, pp. 361–377, Jun 2018, doi: 10.1007/s10899-017-9703-5.

[13] R. Rodriguez-Monguio, M. Errea, and R. Volberg, “Comorbid pathological gambling, mental health, and substance use disorders: Health-care services provision by clinician specialty,” J Behav Addict, vol. 6, no. 3, pp. 406–415, Sep 1 2017, doi: 10.1556/2006.6.2017.054.

[14] S. Cowlishaw, J. K. Hakes, and N. A. Dowling, “Gambling problems in treatment for affective disorders: Results from the National Epidemiologic Survey on Alcohol and Related Conditions (NESARC),” J Affect Disord, vol. 202, pp. 110–4, Sep 15 2016, doi: 10.1016/j.jad.2016.05.023.

[15] D. M. Ledgerwood, M. A. Steinberg, R. Wu, and M. N. Potenza, “Self-reported gambling-related suicidality among gambling helpline callers,” Psychol Addict Behav, vol. 19, no. 2, pp. 175–183, Jun 2005, doi: 10.1037/0893-164X.19.2.175.

[16] N. M. Petry and B. D. Kiluk, “Suicidal ideation and suicide attempts in treatment-seeking pathological gamblers,” J Nerv Ment Dis, vol. 190, no. 7, pp. 462–9, Jul 2002, doi: 10.1097/00005053-200207000-00007.

[17] S. Cox and National Council on Problem Gambling. Gambling, Problem and Pathological Gambling in America: The National Picture. National Council on Problem Gambling, 1997.

[18] R. Etuk, S. D. Shirk, J. Grubbs, and S. W. Kraus, “Gambling problems in US military veterans,” Current Addiction Reports, vol. 7, pp. 210–228, 2020.

[19] P. Nieberg, “Gambling addiction in the military may be going unnoticed, advocates warn,” Task and Purpose, 3 July 2025. [Online]. Available: https://taskandpurpose.com/military-life/research-on-military-gambling-disorders/ (accessed 30 Oct 2026).

[20] National Council on Problem Gambling. “Gambling Addiction Prevention (GAP) Act.” https://www.ncpgambling.org/advocacy/gap-act/#:∼:text=Why%20the%20GAP%20Act%20Matters,stigma%20associated%20with%20the%20disorder (accessed 30 Oct 2026).

[21] (2017). GAO-17-114, Military Personnel: DOD and the Coast Guard Need to Screen for Gambling Disorder Addiction and Update Guidance. [Online] Available: https://www.gao.gov/assets/gao-17-114.pdf (accessed 30 Oct 2026).

[22] (2025). DOD INSTRUCTION 1010.04 PROBLEMATIC SUBSTANCE USE AND GAMBLING DISORDER [Online] Available: https://www.esd.whs.mil/Portals/54/Documents/DD/issuances/dodi/101004p.pdf (accessed 30 Oct 2026).

[23] J. Westermeyer, J. Canive, P. Thuras, M. Oakes, and M. Spring, “Pathological and problem gambling among veterans in clinical care: prevalence, demography, and clinical correlates,” Am J Addict, vol. 22, no. 3, pp. 218–25, May-Jun 2013, doi: 10.1111/j.1521-0391.2012.12011.x.

[24] A. N. Daghestani, E. Elenz, and J. W. Crayton, “Pathological gambling in hospitalized substance abusing veterans,” J Clin Psychiatry, vol. 57, no. 8, pp. 360–3, Aug 1996. [Online]. Available: https://www.ncbi.nlm.nih.gov/pubmed/8752019 (accessed 30 Oct 2026).

[25] S. D. Shirk et al., “Gambling-related cognitive distortions predict level of function among US veterans seeking treatment for gambling disorders,” Am J Addict, vol. 27, no. 2, pp. 108–115, Mar 2018, doi: 10.1111/ajad.12685.

[26] M. N. Potenza, H. Xian, K. Shah, J. F. Scherrer, and S. A. Eisen, “Shared genetic contributions to pathological gambling and major depression in men,” Arch Gen Psychiatry, vol. 62, no. 9, pp. 1015–21, Sep 2005, doi: 10.1001/archpsyc.62.9.1015.

[27] J. F. Scherrer, H. Xian, W. S. Slutske, S. A. Eisen, and M. N. Potenza, “Associations between obsessive-compulsive classes and pathological gambling in a national cohort of male twins,” JAMA Psychiatry, vol. 72, no. 4, pp. 342–9, Apr 2015, doi: 10.1001/jamapsychiatry.2014.2497.

[28] W. S. Slutske et al., “A twin study of the association between pathological gambling and antisocial personality disorder,” J Abnorm Psychol, vol. 110, no. 2, pp. 297–308, May 2001, doi: 10.1037//0021-843x.110.2.297.

[29] R. C. Kessler et al., “DSM-IV pathological gambling in the National Comorbidity Survey Replication,” Psychol Med, vol. 38, no. 9, pp. 1351–60, Sep 2008, doi: 10.1017/S0033291708002900.

[30] S. W. Whiting, M. N. Potenza, C. L. Park, S. A. McKee, C. M. Mazure, and R. A. Hoff, “Investigating Veterans’ Pre-, Peri-, and Post-Deployment Experiences as Potential Risk Factors for Problem Gambling,” J Behav Addict, vol. 5, no. 2, pp. 213–20, Jun 2016, doi: 10.1556/2006.5.2016.027.

[31] I. Parhami, R. Mojtabai, R. J. Rosenthal, T. O. Afifi, and T. W. Fong, “Gambling and the onset of comorbid mental disorders: a longitudinal study evaluating severity and specific symptoms,” J Psychiatr Pract, vol. 20, no. 3, pp. 207–19, May 2014, doi: 10.1097/01.pra.0000450320.98988.7c.

[32] S. Ronzitti, S. W. Kraus, S. E. Decker, and L. Ashrafioun, “Clinical characteristics of veterans with gambling disorders seeking pain treatment,” Addict Behav, vol. 95, pp. 160–165, Aug 2019, doi: 10.1016/j.addbeh.2019.03.014.

[33] O. Kausch, “Suicide attempts among veterans seeking treatment for pathological gambling,” J Clin Psychiatry, vol. 64, no. 9, pp. 1031–8, Sep 2003, doi: 10.4088/jcp.v64n0908.

[34] O. Kausch, L. Rugle, and D. Y. Rowland, “Lifetime histories of trauma among pathological gamblers,” Am J Addict, vol. 15, no. 1, pp. 35–43, Jan-Feb 2006, doi: 10.1080/10550490500419045.

[35] E. A. Stefanovics, M. N. Potenza, J. Tsai, and R. H. Pietrzak, “Prevalence and Clinical Characteristics of Recreational and At-Risk/Problematic Gambling in a National Sample of U.S. Military Veterans,” J Gambl Stud, vol. 39, no. 3, pp. 1077–1097, Sep 2023, doi: 10.1007/s10899-022-10165-z.

[36] L. Blank, S. Baxter, H. B. Woods, and E. Goyder, “Should screening for risk of gambling-related harm be undertaken in health, care and support settings? A systematic review of the international evidence,” Addict Sci Clin Pract, vol. 16, no. 1, p. 35, May 29 2021, doi: 10.1186/s13722-021-00243-9.

[37] T. E. Workman et al., “A Comparison of Veterans with Problematic Opioid Use Identified through Natural Language Processing of Clinical Notes versus Using Diagnostic Codes,” Healthcare (Basel), vol. 12, no. 7, Apr 6 2024, doi: 10.3390/healthcare12070799.

[38] A. P. Association, Diagnostic and Statistical Manual of Mental Disorders, 5th ed, text rev. ed. American Psychiatric Association Publishing, 2022.

